## Supplement for "Functional improvement is a better predictor of steady work than medical improvement for individuals with mental health conditions"

April 17, 2025

#### 1 Supplemental Methods

We model the probability of steady work for participant $n$ in year $j$ using the likelihood

$$y_{n,j} \sim \mathrm{Bern}(\sigma(\mu_n)) \tag{1}$$
$$\mu_{n,j} = \alpha_n + \mathbf{x}_{n,j}\boldsymbol{\beta} \tag{2}$$

where $\sigma$ is the sigmoid function, and $\mathbf{x}_{n,j} \in \mathbb{R}^p$ is the associated vector of predictors of dimension $p$. In this formulation, the person-specific parameters $\alpha_n$ are random intercept terms.

##### 1.1 Missingness

We used marginalization in order to resolve missingness in the dataset. We first determined whether each predictor variable is a count, or otherwise real-valued. We then fit predictive models to each predictor variable, simultaneously for integer count-based predictors

$$x_{n,j,y} \sim \mathrm{Poisson}(r_{n,j,y}) \qquad r_{n,j} = \gamma_{n,j} + \sum_{k\neq j} \eta_k x_{n,k,y},$$

and otherwise real-valued predictors

$$x_{n,j,y} \sim \mathrm{Normal}(\sigma(r_{m,j,y}), \sigma_j) \qquad r_{n,j,y} = \gamma n, j + \sum_{k\neq j} \eta_k x_{n,k,y},$$

for each variable $j$, year $y$, and person $n$. In essence we model each unknown value for a given year as a generalized linear regression on the other values for that year and a person-specific random effect that is pooled across years.

In sampling the full model's posterior distribution and computing the statistics for $\beta$ and $\alpha_n$, we marginalize over all of the model parameters for the missing observations.

##### 1.2 Bayesian model priors

We regularize the model by using a combination of weakly informative and sparsity promoting priors,

$$\begin{aligned} \alpha_n &\sim \mathrm{normal}(0, 10) \\ \boldsymbol{\beta} &\sim \text{Finnish-Horseshoe}(n), \end{aligned} \tag{3}$$

where the Finnish horseshoe prior [Piironen and Vehtari, 2016, 2017b] promotes sparsity-based regularization on $\boldsymbol{\beta}$.

#### 2 Supplemental Results

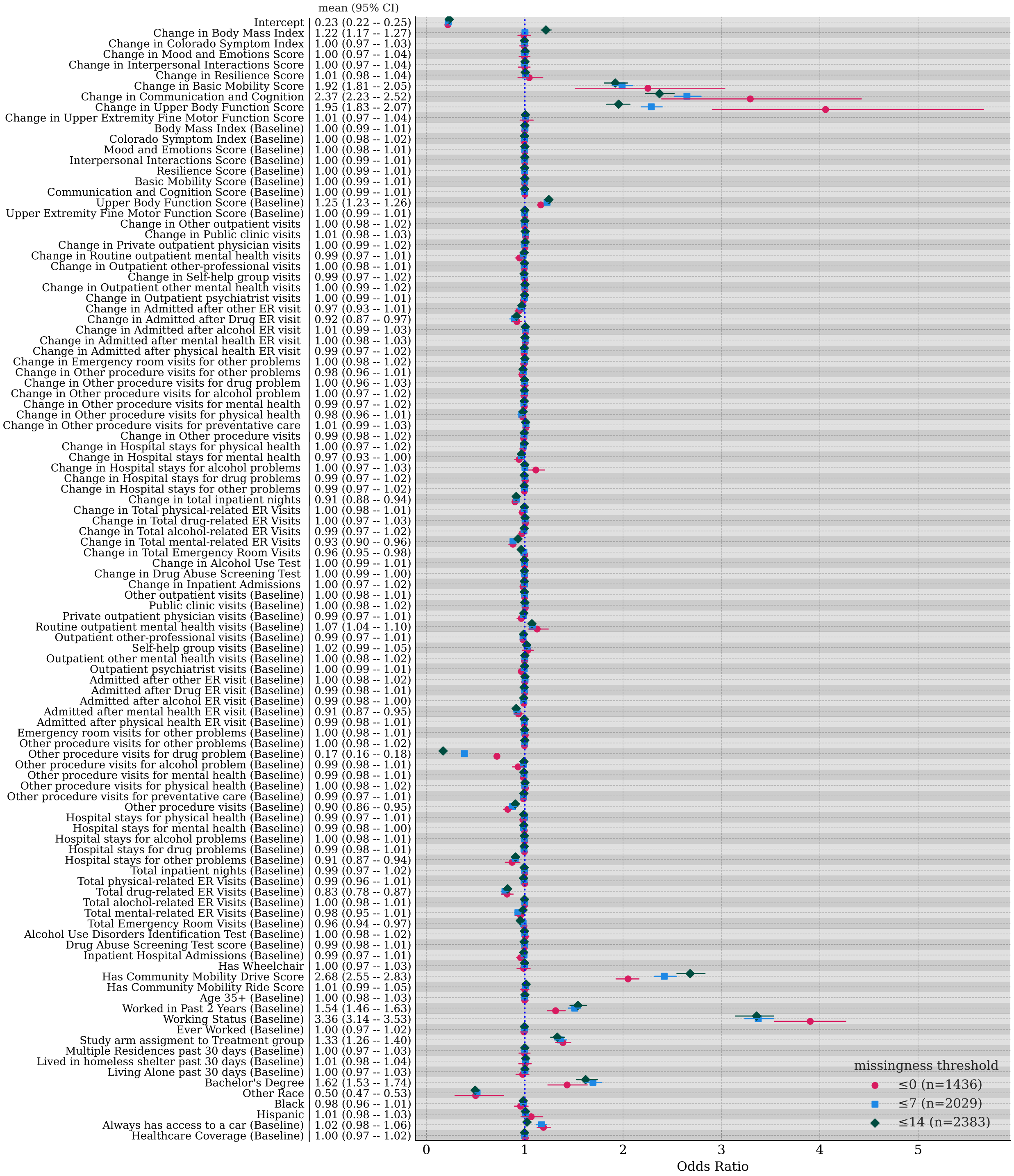


Figure 1: Comparing models based on missingness thresholds, finding that the models were largely consistent in terms of findings.

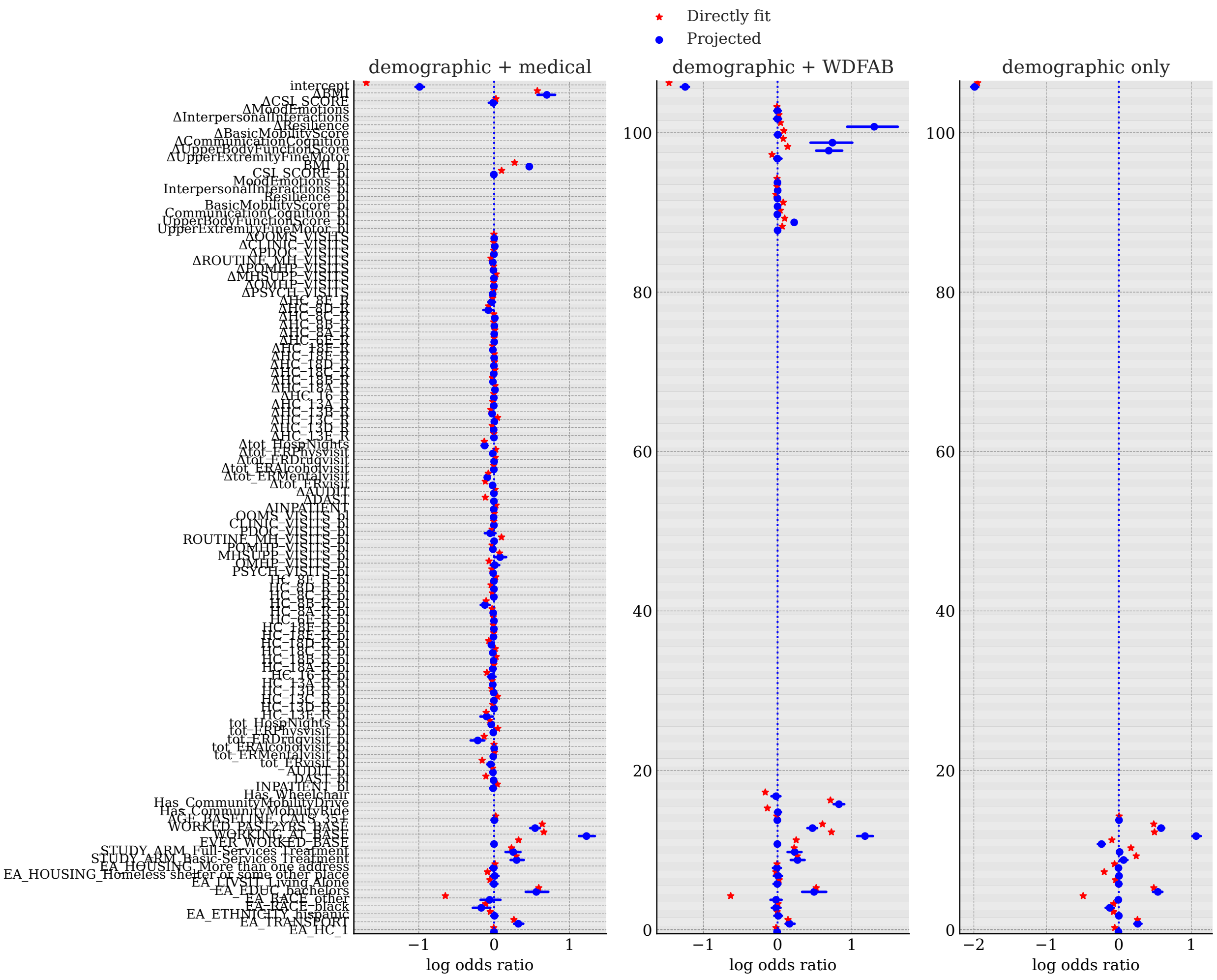


Figure 2: Regression coefficients for submodels fitted either directly or through a posterior predictive projection method [Catalina et al., 2022, Pavone et al., 2023, Piironen et al., 2020, Piironen and Vehtari, 2017a, Weber et al., 2025]

### 3 Source code

The following code describes the model

```

from collections import defaultdict
from bayesianquilts import BayesianModel
import tensorflow as tf
from tensorflow_probability.python import distributions as tfd
from tensorflow_probability.python import bijectors as tfb
from bayesianquilts.vi.advi import build_surrogate_posterior
from bayesianquilts.util import batched_minimize

import numpy as np
import pandas as pd
from sklearn import metrics
from psisloo import psisloo
from tqdm import tqdm
import sys

from nppsis import psisloo, psislw

# mu0 is person/variable specific
# mu00 is variable specific


class ImputedBernModelRescaled(BayesianModel):
    def __init__(
        self,
        n_people,
        gaussian_yearly_vars,
        poisson_yearly_vars,
        binary_yearly_vars,
        baseline_vars,
        outcome_var=None,
        outcome_tau=1.0,
        beta_outcome_scale=2.0,
        init=None,
        means=None,
        stds=None,
        diff_means=None,
        diff_stds=None,
        dtype=tf.float64,
    ):
        if outcome_var is None:
            outcome_var = "STEADY_WORKER"
        self.outcome_var = outcome_var
        self.outcome_tau = outcome_tau
        self.dtype = dtype
        self.strategy = None
        self.gaussian_yearly_vars = gaussian_yearly_vars
        self.diff_means = diff_means if diff_means is not None else defaultdict(float)
        self.diff_stds = diff_stds if diff_stds is not None else defaultdict(lambda: 1)
        self.means = means if means is not None else defaultdict(float)
        self.stds = stds if stds is not None else defaultdict(lambda: 1)
        self.gaussian_yearly_vars_loc = tf.cast(
            [self.means.get(c, 0) for c in gaussian_yearly_vars], self.dtype
        )
        self.gaussian_yearly_vars_scale = tf.cast(
            [self.stds.get(c, 1) for c in gaussian_yearly_vars], self.dtype
        )
        self.gaussian_yearly_vars_diff_loc = tf.cast(
            [self.diff_means.get(c, 0) for c in gaussian_yearly_vars], self.dtype
        )
        self.gaussian_yearly_vars_diff_scale = tf.cast(
            [self.diff_stds.get(c, 1) for c in gaussian_yearly_vars], self.dtype
        )
        self.binary_yearly_vars = binary_yearly_vars
        self.poisson_yearly_vars = poisson_yearly_vars
        self.poisson_yearly_vars_loc = tf.cast(
            [self.means.get(c, 0) for c in poisson_yearly_vars], self.dtype
        )
        self.poisson_yearly_vars_scale = tf.cast(
            [self.stds.get(c, 1) for c in poisson_yearly_vars], self.dtype
        )
```

```python
        self.poisson_yearly_vars_diff_loc = tf.cast(
            [self.diff_means.get(c, 0) for c in poisson_yearly_vars], self.dtype
        )
        self.poisson_yearly_vars_diff_scale = tf.cast(
            [self.diff_stds.get(c, 1) for c in poisson_yearly_vars], self.dtype
        )
        self.baseline_vars = baseline_vars
        self.beta_outcome_scale = beta_outcome_scale
        self.n_people = n_people
        self.init = init

        self.create_distributions(init=init)

    def create_distributions(self, init=None):
        self.prior_distribution = tfd.JointDistributionNamed(
            {
                "alpha_outcome": tfd.Independent(
                    tfd.Normal(
                        loc=tf.zeros([1], self.dtype),
                        scale=10 * tf.ones([1], self.dtype),
                    ),
                    reinterpreted_batch_ndims=1,
                ),
                "mu0_gaussian": tfd.Independent(
                    tfd.Normal(
                        loc=tf.zeros(
                            [self.n_people, len(self.gaussian_yearly_vars), 1],
                            dtype=self.dtype,
                        ),
                        scale=3
                        * tf.ones(
                            [self.n_people, len(self.gaussian_yearly_vars), 1],
                            dtype=self.dtype,
                        ),
                    ),
                    reinterpreted_batch_ndims=3,
                ),
                "mu0_poisson": tfd.Independent(
                    tfd.Normal(
                        loc=tf.zeros(
                            [self.n_people, len(self.poisson_yearly_vars), 1],
                            dtype=self.dtype,
                        ),
                        scale=3
                        * tf.ones(
                            [self.n_people, len(self.poisson_yearly_vars), 1],
                            dtype=self.dtype,
                        ),
                    ),
                    reinterpreted_batch_ndims=3,
                ),
                "mu00_gaussian": tfd.Independent(
                    tfd.Normal(
                        loc=tf.zeros(
                            [1, len(self.gaussian_yearly_vars), 1], dtype=self.dtype
                        ),
                        scale=6
                        * tf.ones(
                            [1, len(self.gaussian_yearly_vars), 1], dtype=self.dtype
                        ),
                    ),
                    reinterpreted_batch_ndims=3,
                ),
                "mu00_poisson": tfd.Independent(
                    tfd.Normal(
                        loc=tf.zeros(
                            [1, len(self.poisson_yearly_vars), 1], dtype=self.dtype
                        ),
                        scale=5
                        * tf.ones(
                            [1, len(self.poisson_yearly_vars), 1], dtype=self.dtype
                        ),
                    ),
                    reinterpreted_batch_ndims=3,
                ),
                "sd00_gaussian": tfd.Independent(
```

```
                    tfd.HalfNormal(
                        scale=tf.ones(
                            [1, len(self.gaussian_yearly_vars), 1], dtype=self.dtype
                        ),
                    ),
                    reinterpreted_batch_ndims=3,
                ),
                "beta_outcome": tfd.Independent(
                    tfd.Horseshoe(
                        # loc=tf.zeros(
                        #     [
                        #         len(baseline_vars + has_vars)
                        #         + len(poisson_yearly_vars) * 2
                        #         + len(gaussian_yearly_vars) * 2
                        #     ],
                        #     self.dtype,
                        # ),
                        scale=self.beta_outcome_scale
                        / np.sqrt(3 * self.n_people)
                        * tf.ones(
                            [
                                len(self.baseline_vars)
                                + len(self.binary_yearly_vars)
                                + len(self.poisson_yearly_vars) * 2
                                + len(self.gaussian_yearly_vars) * 2
                            ],
                            self.dtype,
                        ),
                    ),
                    reinterpreted_batch_ndims=1,
                ),
                "beta_gaussian": tfd.Independent(
                    tfd.Normal(
                        loc=tf.zeros(
                            [
                                len(self.baseline_vars)
                                + len(self.binary_yearly_vars)
                                + len(self.poisson_yearly_vars),
                                len(self.gaussian_yearly_vars),
                            ],
                            self.dtype,
                        ),
                        scale=self.outcome_tau
                        * tf.ones(
                            [
                                len(self.baseline_vars)
                                + len(self.binary_yearly_vars)
                                + len(self.poisson_yearly_vars),
                                len(self.gaussian_yearly_vars),
                            ],
                            self.dtype,
                        ),
                    ),
                    reinterpreted_batch_ndims=2,
                ),
                "beta_poisson": tfd.Independent(
                    tfd.Normal(
                        loc=tf.zeros(
                            [
                                len(self.baseline_vars)
                                + len(self.binary_yearly_vars)
                                + len(self.gaussian_yearly_vars),
                                len(self.poisson_yearly_vars),
                            ],
                            self.dtype,
                        ),
                        scale=self.outcome_tau
                        * tf.ones(
                            [
                                len(self.baseline_vars)
                                + len(self.binary_yearly_vars)
                                + len(self.gaussian_yearly_vars),
                                len(self.poisson_yearly_vars),
                            ],
                            self.dtype,
                        ),
```

```
                    ),
                    reinterpreted_batch_ndims=2,
                ),
            }
        )
        bijectors = defaultdict(lambda: tfb.Identity())
        bijectors["sd0_gaussian"] = tfb.Softplus()
        bijectors["sd00_gaussian"] = tfb.Softplus()
        initializers = {
            "beta_gaussian": tf.zeros(
                [
                    len(self.baseline_vars)
                    + len(self.binary_yearly_vars)
                    + len(self.poisson_yearly_vars),
                    len(self.gaussian_yearly_vars),
                ],
                self.dtype,
            ),
            "beta_poisson": tf.zeros(
                [
                    len(self.baseline_vars)
                    + len(self.binary_yearly_vars)
                    + len(self.gaussian_yearly_vars),
                    len(self.poisson_yearly_vars),
                ],
                self.dtype,
            ),
            "beta_outcome": tf.zeros(
                [
                    len(self.baseline_vars)
                    + len(self.binary_yearly_vars)
                    + len(self.poisson_yearly_vars) * 2
                    + len(self.gaussian_yearly_vars) * 2
                ],
                self.dtype,
            ),
            "mu0_gaussian": tf.zeros(
                [self.n_people, len(self.gaussian_yearly_vars), 1], dtype=self.dtype
            ),
            "mu0_poisson": tf.zeros(
                [self.n_people, len(self.poisson_yearly_vars), 1], dtype=self.dtype
            ),
            "sd00_gaussian": tf.ones(
                [1, len(self.gaussian_yearly_vars), 1], dtype=self.dtype
            ),
        }
        self.beta_outcome_labels = (
            self.baseline_vars
            + self.binary_yearly_vars
            + [f"{c}_bl" for c in self.poisson_yearly_vars]
            + [f"{c}" for c in self.poisson_yearly_vars]
            + [f"{c}_bl" for c in self.gaussian_yearly_vars]
            + [f"{c}" for c in self.gaussian_yearly_vars]
        )
        if init is not None:
            test = self.prior_distribution.sample()
            to_delete = []
            for k, v in init.items():
                if np.array_equiv(test[k].shape.as_list(), v.shape.as_list()):
                    print(f"Using prior {k}", flush=True)
                    initializers[k] = v
                else:
                    to_delete += [k]
            for k in to_delete:
                del self.init[k]
        self.surrogate_distribution = build_surrogate_posterior(
            self.prior_distribution, bijectors=bijectors, initializers=initializers
        )
        return

    def index_persons(self, param, person):
        shape = param.shape.as_list()
        trans = [1, 0] + list(range(2, len(shape)))
        param = tf.transpose(param, trans)
        param = tf.gather(param, person)
        param = tf.transpose(param, trans)
```

```
        return param

    def predictive_distribution(self, data, **params):
        if "_preprocessed" not in data.keys():
            data = self.preprocess(data)
        mu0_gaussian = params["mu0_gaussian"]
        mu00_gaussian = params["mu00_gaussian"]
        beta_gaussian = params["beta_gaussian"]
        beta_poisson = params["beta_poisson"]
        beta_outcome = params["beta_outcome"]
        alpha_outcome = params["alpha_outcome"]

        mu0_poisson = params["mu0_poisson"]
        mu00_poisson = params["mu00_poisson"]

        sd = params["sd00_gaussian"]
        person = data["person_id"]
        # sd = self.index_persons(sd, person)

        mu0_gaussian = self.index_persons(mu0_gaussian, person)

        mu0_poisson = self.index_persons(mu0_poisson, person)

        mu_gaussian = mu0_gaussian + mu00_gaussian
        mu_poisson = mu0_poisson + mu00_poisson

        if len(self.gaussian_yearly_vars) > 0:
            gaussian_yearly = (
                tf.cast(data["gaussian_yearly"], self.dtype)
                - self.gaussian_yearly_vars_diff_loc[:, tf.newaxis]
            ) / self.gaussian_yearly_vars_scale[:, tf.newaxis]
            # plug-in using imputation model
            gaussian_yearly_ = gaussian_yearly + tf.zeros_like(mu_gaussian)
            gaussian_yearly_ = tf.where(
                tf.math.is_finite(gaussian_yearly_), gaussian_yearly_, mu_gaussian
            )
        else:
            gaussian_yearly_ = 0
        if len(self.poisson_yearly_vars) > 0:
            poisson_yearly = data["poisson_yearly"]
            poisson_yearly_ = poisson_yearly + tf.zeros_like(mu_poisson)
            poisson_yearly_ = tf.where(
                tf.math.is_finite(poisson_yearly_),
                poisson_yearly_,
                tf.math.exp(mu_poisson),
            )
        else:
            poisson_yearly_ = 0

        if len(self.baseline_vars) > 0:
            baseline = tf.cast(data["baseline_vars"], self.dtype)
            baseline = baseline[tf.newaxis, ..., tf.newaxis]
        else:
            baseline = 0

        if len(self.binary_yearly_vars) > 0:
            has_yearly = tf.cast(data["binary_yearly"], self.dtype)
        else:
            has_yearly = 0

        # regressors
        ## gaussian yearly predictors

        gaussian_ea = tf.reduce_sum(
            beta_gaussian[:, tf.newaxis, : len(self.baseline_vars), :] * baseline,
            axis=-2,
        )[
            ..., tf.newaxis
        ]  # batch x person x n_gaussian x year (1)
        if len(self.binary_yearly_vars) > 0:
            gaussian_has = tf.reduce_sum(
                beta_gaussian[
                    :,
                    tf.newaxis,
                    len(self.baseline_vars) : len(
                        self.baseline_vars + self.binary_yearly_vars
```

```
                    ),
                ][..., tf.newaxis]
                * has_yearly[..., tf.newaxis, :],
                axis=-3,
            )
            poisson_has = tf.reduce_sum(
                beta_poisson[
                    :,
                    tf.newaxis,
                    len(self.baseline_vars) : len(
                        self.baseline_vars + self.binary_yearly_vars
                    ),
                ][..., tf.newaxis]
                * has_yearly[..., tf.newaxis, :],
                axis=-3,
            )
            outcome_has = tf.reduce_sum(
                beta_outcome[
                    ...,
                    tf.newaxis,
                    len(self.baseline_vars) : len(
                        self.baseline_vars + self.binary_yearly_vars
                    ),
                    tf.newaxis,
                ]
                * has_yearly[..., 1:],
                axis=-2,
            )
        else:
            gaussian_has = 0
            poisson_has = 0
            outcome_has = 0

        if len(self.poisson_yearly_vars) > 0:
            gaussian_poisson = tf.reduce_sum(
                beta_gaussian[
                    :, tf.newaxis, len(self.baseline_vars + self.binary_yearly_vars) :
                ][..., tf.newaxis]
                * (
                    poisson_yearly_[..., tf.newaxis, :]
                    - self.poisson_yearly_vars_loc[:, tf.newaxis, tf.newaxis]
                )
                / self.poisson_yearly_vars_scale[:, tf.newaxis, tf.newaxis],
                axis=-3,
            )
            outcome_poisson = tf.reduce_sum(
                beta_outcome[
                    ...,
                    tf.newaxis,
                    len(self.baseline_vars + self.binary_yearly_vars) : len(
                        self.baseline_vars
                        + self.binary_yearly_vars
                        + self.poisson_yearly_vars
                    ),
                    tf.newaxis,
                ]
                * (
                    poisson_yearly_[..., 0:1]
                    - self.poisson_yearly_vars_loc[:, tf.newaxis]
                )
                / self.poisson_yearly_vars_scale[:, tf.newaxis],
                axis=-2,
            )

            delta_poisson_yearly = poisson_yearly_[..., 1:] - poisson_yearly_[..., 0:1]
            outcome_poisson_delta = tf.reduce_sum(
                beta_outcome[
                    ...,
                    tf.newaxis,
                    len(
                        self.baseline_vars
                        + self.binary_yearly_vars
                        + self.poisson_yearly_vars
                    ) : len(
                        self.baseline_vars
                        + self.binary_yearly_vars
```

```
                        + self.poisson_yearly_vars
                        + self.poisson_yearly_vars
                    ),
                    tf.newaxis,
                ]
                * (delta_poisson_yearly - self.poisson_yearly_vars_loc[:, tf.newaxis])
                / self.poisson_yearly_vars_scale[:, tf.newaxis],
                axis=-2,
            )

        else:
            gaussian_poisson = 0
            outcome_poisson = 0
            outcome_poisson_delta = 0

        mu_gaussian += gaussian_ea + gaussian_has + gaussian_poisson

        ## poisson yearly predictors

        poisson_ea = tf.reduce_sum(
            beta_poisson[:, tf.newaxis, : len(self.baseline_vars), :] * baseline,
            axis=-2,
        )[
            ..., tf.newaxis
        ]  # batch x person x n_gaussian x year (1)

        if len(self.gaussian_yearly_vars) > 0:
            poisson_gaussian = tf.reduce_sum(
                beta_poisson[
                    :, tf.newaxis, len(self.baseline_vars + self.binary_yearly_vars) :
                ][..., tf.newaxis]
                * (
                    gaussian_yearly_[..., tf.newaxis, :]-self.gaussian_yearly_vars_loc[:, tf.newaxis, tf.newaxis]
                )
                / self.gaussian_yearly_vars_scale[:, tf.newaxis, tf.newaxis],
                axis=-3,
            )
            outcome_gaussian = tf.reduce_sum(
                beta_outcome[
                    ...,
                    tf.newaxis,
                    len(
                        self.baseline_vars
                        + self.binary_yearly_vars
                        + self.poisson_yearly_vars
                        + self.poisson_yearly_vars
                    ) : len(
                        self.baseline_vars
                        + self.binary_yearly_vars
                        + self.poisson_yearly_vars
                        + self.poisson_yearly_vars
                        + self.gaussian_yearly_vars
                    ),
                    tf.newaxis,
                ]
                * (
                    gaussian_yearly_[..., 0:1]
                    - self.gaussian_yearly_vars_loc[:, tf.newaxis]
                )
                / self.gaussian_yearly_vars_scale[:, tf.newaxis],
                axis=-2,
            )
            delta_gaussian_yearly = (
                gaussian_yearly_[..., 1:] - gaussian_yearly_[..., 0:1]
            )
            outcome_gaussian_delta = tf.reduce_sum(
                beta_outcome[
                    ...,
                    tf.newaxis,
                    len(
                        self.baseline_vars
                        + self.binary_yearly_vars
                        + self.poisson_yearly_vars
                        + self.poisson_yearly_vars
                        + self.gaussian_yearly_vars
                    ) :,
```

```python
                    tf.newaxis,
                ]
                * (
                    delta_gaussian_yearly
                    - self.gaussian_yearly_vars_diff_loc[:, tf.newaxis]
                )
                / self.gaussian_yearly_vars_diff_scale[:, tf.newaxis],
                axis=-2,
            )
        else:
            poisson_gaussian = 0
            outcome_gaussian = 0
            outcome_gaussian_delta = 0

        mu_poisson += (
            poisson_ea
            + poisson_has
            + poisson_gaussian
            - tf.math.log(self.poisson_yearly_vars_scale)[..., tf.newaxis]
        )

        ## outcome
        outcome_ea = tf.reduce_sum(
            beta_outcome[..., tf.newaxis, : len(self.baseline_vars), tf.newaxis]
            * baseline,
            axis=-2,
        )

        # model the values
        if len(self.gaussian_yearly_vars) > 0:
            rv_gaussian = tfd.Normal(mu_gaussian, sd)
            ll_gaussian = rv_gaussian.log_prob(gaussian_yearly)
            ll_gaussian = tf.where(
                tf.math.is_finite(ll_gaussian), ll_gaussian, tf.zeros_like(ll_gaussian)
            )
            # ll_gaussian *= tf.cast(tf.math.is_finite(gaussian_yearly), self.dtype)
            ll_gaussian = tf.reduce_mean(ll_gaussian, axis=[-1, -2])
        else:
            ll_gaussian = 0
        if len(self.poisson_yearly_vars) > 0:
            rv_poisson = tfd.Poisson(log_rate=mu_poisson)
            ll_poisson = rv_poisson.log_prob(poisson_yearly)
            ll_poisson = tf.where(
                tf.math.is_finite(ll_poisson), ll_poisson, tf.zeros_like(ll_poisson)
            )
            ll_poisson *= tf.cast(tf.math.is_finite(poisson_yearly), self.dtype)
            ll_poisson = tf.reduce_mean(ll_poisson, axis=[-1, -2])
        else:
            ll_poisson = 0

        # model the outcome
        mu_outcome = (
            outcome_gaussian
            + outcome_ea
            + outcome_poisson
            + outcome_has
            + alpha_outcome[..., tf.newaxis]
            + outcome_gaussian_delta
            + outcome_poisson_delta
        )
        outcome = data[self.outcome_var][..., 1:]
        mu_outcome += tf.zeros_like(outcome)
        rv_outcome = tfd.Bernoulli(logits=mu_outcome)
        ll_outcome = rv_outcome.log_prob(outcome)
        ll = ll_gaussian + ll_poisson + tf.reduce_sum(ll_outcome, axis=-1)
        return {
            "log_likelihood": ll,
            "ll_outcome": ll_outcome,
            "ll_poisson": ll_poisson,
            "ll_gaussian": ll_gaussian,
            "rv_outcome": rv_outcome,
        }

    def log_likelihood(self, data, **params):
        return self.predictive_distribution(data, **params)["log_likelihood"]

```

```python
    def unormalized_log_prob(self, data=None, prior_weight=tf.constant(1.0), **params):
        prediction = self.predictive_distribution(data, **params)
        log_likelihood = prediction["log_likelihood"]
        max_val = tf.reduce_max(log_likelihood)

        finite_portion = tf.where(
            tf.math.is_finite(log_likelihood),
            log_likelihood,
            tf.zeros_like(log_likelihood),
        )
        min_val = tf.reduce_min(finite_portion) - 1.0
        log_likelihood = tf.where(
            tf.math.is_finite(log_likelihood),
            log_likelihood,
            tf.ones_like(log_likelihood) * min_val,
        )
        prior = self.prior_distribution.log_prob(params)
        prior_weight = tf.cast(prior_weight, self.dtype)
        return tf.reduce_sum(log_likelihood, axis=-1) + prior_weight * prior

    def reverse_kl(self, data, model, **params):
        prediction = self.predictive_distribution(data, **params)["rv_outcome"]
        other_prediction = model.predictive_distribution(data, **params)["rv_outcome"]
        kl = other_prediction.kl_divergence(prediction)
        return kl

    def fit_projection(
        self, other, batched_data_factory, num_steps, samples=32, **kwargs
    ):
        def objective(data):
            this_prediction = self.predictive_distribution(
                data, **self.sample(samples)
            )["rv_outcome"]
            other_prediction = other.predictive_distribution(
                data, **other.sample(samples)
            )["rv_outcome"]
            delta = other_prediction.kl_divergence(this_prediction)
            return tf.reduce_mean(delta)

        return batched_minimize(
            objective,
            batched_data_factory=batched_data_factory,
            num_steps=num_steps,
            trainable_variables=self.surrogate_distribution.variables,
            **kwargs,
        )

    def preprocess(self, record):
        out = {
            self.outcome_var: record[self.outcome_var],
            "person_id": record["person_id"],
        }
        if len(self.gaussian_yearly_vars) > 0:
            out["gaussian_yearly"] = tf.concat(
                [
                    tf.cast(record[c][..., tf.newaxis, :], self.dtype)
                    for c in self.gaussian_yearly_vars
                ],
                axis=-2,
            )
        if len(self.poisson_yearly_vars) > 0:
            out["poisson_yearly"] = tf.concat(
                [
                    tf.cast(record[c][:, tf.newaxis], self.dtype)
                    for c in self.poisson_yearly_vars
                ],
                axis=-2,
            )
        if len(self.baseline_vars) > 0:
            out["baseline_vars"] = tf.concat(
                [
                    tf.cast(record[c][:, tf.newaxis], self.dtype)
                    for c in self.baseline_vars
                ],
                axis=-1,
            )
```

```
        if len(self.binary_yearly_vars) > 0:
            out["binary_yearly"] = tf.concat(
                [
                    tf.cast(record[c][:, tf.newaxis], self.dtype)
                    for c in self.binary_yearly_vars
                ],
                axis=-2,
            )
        return out

    def loo_auc(self, data_factory, samples=32):
        ll = []
        labels = []
        predicted = []
        _p = self.sample(samples)
        for batch in tqdm(iter(data_factory())):
            pred = self.predictive_distribution(batch, **_p)
            ll += [pred["ll_outcome"]]
            predicted += [pred["rv_outcome"].prob(1.0)]
            labels += [batch[self.outcome_var][:, 1:]]
        ll = tf.concat(ll, axis=1)
        predicted = tf.concat(predicted, axis=1)

        labels = tf.concat(labels, axis=0)
        finite_portion = tf.where(
            tf.math.is_finite(ll),
            ll,
            tf.zeros_like(ll),
        )
        min_val = tf.reduce_min(finite_portion) - 1.0
        ll = tf.where(
            tf.math.is_finite(ll),
            ll,
            tf.ones_like(ll) * min_val,
        )

        lw, khat = psislw(-tf.reduce_sum(ll, axis=-1).numpy())
        w = tf.math.exp(lw)
        w = (w / tf.reduce_sum(w, axis=0, keepdims=True))[..., tf.newaxis]

        psis_pred_y = tf.reduce_sum(predicted * w, axis=0)
        loo = psisloo(tf.reduce_sum(ll, axis=-1).numpy())
        print(f"loo: {loo}")

        predicted = tf.reduce_mean(predicted, axis=0)
        predicted = np.reshape(predicted, -1)
        psis_pred_y = np.reshape(psis_pred_y, -1)
        labels = np.reshape(labels, -1)
        loo_roc = auroc(labels, psis_pred_y)
        loo_prc = auprc(labels, psis_pred_y)

        roc = auroc(labels, predicted)
        prc = auprc(labels, predicted)

        return {
            "loo": loo,
            "loo_roc": loo_roc,
            "loo_prc": loo_prc,
            "prc": prc,
            "roc": roc,
            "khat": khat,
        }


def auroc(labels, probs):
    fpr, tpr, thresholds = metrics.roc_curve(labels, probs, pos_label=1)
    return {
        "auroc": metrics.auc(fpr, tpr),
        "fpr": fpr,
        "tpr": tpr,
        "thresholds": thresholds,
    }


def auprc(labels, probs):
    precision, recall, thresholds = metrics.precision_recall_curve(labels, probs)
```

```
    return {
        "auprc": metrics.auc(recall, precision),
        "precision": precision,
        "recall": recall,
        "thresholds": thresholds,
    }


def classification_metrics(
    data_factory,
    prediction_fn,
    by_vars=None,
    outcome_label="label",
    save_file=None,
):
    if by_vars is None:
        by_vars = []
    collect_vars = by_vars + [outcome_label]
    collect_vars = set(collect_vars)
    collected_data = {k: [] for k in collect_vars}
    probs = []
    metrics = {}

    for batch in iter(data_factory()):
        for k in collected_data.keys():
            collected_data[k] += [tf.squeeze(batch[k]).numpy()]

        probs += [prediction_fn(data=batch)]

    probs = np.concatenate(probs, axis=0)
    for k in collected_data.keys():
        collected_data[k] = np.squeeze(np.concatenate(collected_data[k], axis=0))

    # HACK
    probs = tf.reshape(probs, -1)
    collected_data[outcome_label] = tf.reshape(collected_data[outcome_label][:, 1:], -1)

    computed = pd.DataFrame({"probs": probs, **collected_data})
    if save_file:
        computed.to_parquet(save_file)
    metrics["prob"] = np.mean(computed[outcome_label])
    metrics["auroc"] = auroc(computed[outcome_label], computed.probs)
    metrics["auprc"] = auprc(computed[outcome_label], computed.probs)
    return metrics
```

Juho Piironen and Aki Vehtari. On the Hyperprior Choice for the Global Shrinkage Parameter in the Horseshoe Prior. *arXiv:1610.05559 [stat]*, October 2016.

Juho Piironen and Aki Vehtari. Comparison of Bayesian predictive methods for model selection. *Statistics and Computing*, 27(3):711–735, May 2017a. ISSN 1573-1375. doi: 10.1007/s11222-016-9649-y.

Juho Piironen and Aki Vehtari. Sparsity information and regularization in the horseshoe and other shrinkage priors. *Electronic Journal of Statistics*, 11(2):5018–5051, 2017b. ISSN 1935-7524. doi: 10.1214/17-EJS1337SI.

Juho Piironen, Markus Paasiniemi, and Aki Vehtari. Projective inference in high-dimensional problems: Prediction and feature selection. *Electronic Journal of Statistics*, 14(1):2155–2197, January 2020. ISSN 1935-7524, 1935-7524. doi: 10.1214/20-EJS1711.

Frank Weber, Änne Glass, and Aki Vehtari. Projection predictive variable selection for discrete response families with finite support. *Computational Statistics*, 40(2):701–721, February 2025. ISSN 0943-4062, 1613-9658. doi: 10.1007/s00180-024-01506-0.
